# DECODE the quality of dying in the intensive care unit

**DOI:** 10.1101/2025.07.13.25331477

**Authors:** Sarah Dougan, Graeme J Duke, Stephanie Hunter, Stephanie N Ha, Nicholas J Johnson, David Lehane, Jarel TS Saw, Alistair Teo, Peishan S Zou, Jonathan Barrett

## Abstract

**Objective:** Existing national quality of care processes do not assess quality of care for the dying patient. We developed the Documentation and Evaluation of Care of Dying Equation (DECoDE) score, a 21-point unweighted binary questionnaire. for this purpose.

**Design:** Retrospective clinical audit by clinical staff of medical records.

**Setting:** Two metropolitan adult intensive care units (ICU) in Victoria.

**Participants:** 157 randomly selected records of adults who died in ICU between 1 Jan 2015 – 30 June 2017.

**Main outcomes:** DECoDE audit score, patient characteristics, and therapeutics delivered in the last 24hrs of life.

**Results:** Over 30-months there were 5,194 ICU admissions and 539 (10.4%) deaths - 157 (43.9%) were randomly selected for audit. 122 (82.2%) were receiving curative treatment within the 24-hours prior to death. 130 (83%) deaths were expected and occurred 1.5 (IQR =1-4) days after agreement to withdraw treatment. Median DE-CoDE score was 11 (IQR =9-13) out of a maximum score of 21. Factors associated with a lower DECoDE score included shorter length of stay, unexpected death, and non-oncologic diagnosis. Other patient factors including age, severity of illness (APACHE-IIIJ) score =102 [IQR =80-128]), and organ donation (7%) were not significant.

**Conclusion:** The DECoDE audit tool may be useful for screening of care provided to dying patients in ICU. Not all patients require a high score for high quality care but a low score may indicate poor quality of care, warranting further review.

## Introduction

The quality of intensive care is regularly monitored[1,2] but not the quality of care of the dying.[3,4] While recovery and survival are preferred outcomes following intensive care, for 1 in 10 patients death is unavoidable due to overwhelming illness and/or patient choice.[5,6] On average there is one death in an intensive care unit (ICU) in Australia every hour. Approximately 150,000 adults die in Australia die each year[7] including 10,000 (7%) following admission to (public health sector) ICU.[1,8]

The nature and complexity of critically illness create obstacles to high quality end-of-life care. Diagnosis of dying is problematic and often delayed.[9,10] This limits time available to plan and coordinate high-quality patient-centred end-of-life care.[3,6,11]

Considerable resources have been allocated to improving care of the dying patient in acute healthcare facilities.[6-8,12-14] and monitoring the quality of end-of-life care is essential to achieving this goal. Several different monitoring instruments have been proposed or developed [15–20]. They are, however, labour intensive and impractical outside a research infrastructure and a well-resourced hospital. Most focus on diagnosis and decisions with few elements to measure the care of the dying process.

To our knowledge no audit tool has been developed to assess the quality of care for the dying in ICU, although many of the essential elements are contained within published reports[3–6] and clinical guidelines[11–13] The aim of this project was to develop and test a simple audit tool to assist with monitoring and review of the quality of care of the dying patients and their families. The tool is a simple binary survey tool based on chart review and generates a numerical score - the Documentation and Evaluation of Care of Dying Equation (DECoDE) score.

## Methods

A draft survey tool was derived from elements within published clinical guidelines [12–14]. The desirable characteristics were simplicity, brevity, universality, and clinically relevant. Following an iterative process with clinical staff and community representatives, and a pilot trial, we refined the DECoDE audit tool.

DECoDE contains 21 binary questions (Table 1) that focus on elements of clinical care relevant to the dying patient and their family.[3,4] All elements are relevant to care provided in any ICU or acute health care service. Each element is given a binary score (present or absent) and the cumulative sum informs the final score. Importantly the (crude) score is not risk-adjusted for casemix or local resources.

**Table 1.** DECoDE survey results presented as number (percent) for both hospital sites. P-value comparison between the two sites. LMO = local medical officer; GoC= goals of care; CRP= consensus resuscitation plan. PRN = as required

| No. | Survey Question Description | Site 1,<br>n (%) | Site 2,<br>n (%) | P-value |
| --- | --- | --- | --- | --- |
| 1 | LMO details documented? | 97 (88) | 41 (89) | 1.00 |
| 2 | Emergency contact documented? | 110 (99) | 45 (97) | 0.50 |
| 3 | GoC / CRP documented? | 96 (86) | 40 (87) | 1.00 |
| 4 | SDM/MEPOA documented? | 39 (35) | 16 (35) | 1.00 |
| 5 | “Dying patient” documented? | 99 (89) | 41 (89) | 1.00 |
| 6 | Discussion with patient and/or family? | 100 (91) | 43 (93) | 0.51 |
| 7 | Patient wishes documented? | 9 (8) | 8 (20) | 0.10 |
| 8 | Spiritual care support offered? | 26 (23) | 8 (18) | 0.52 |
| 9 | Social Work referral? | 43 (39) | 26 (57) | 0.05 |
| 10 | Palliative care referral? | 17 (15) | 13 (28) | 0.08 |
| 11 | Appropriate venue discussed? | 7 (6) | 9 (20) | 0.02 |
| 12 | Family present at death? | 89 (80) | 35 (76) | 0.67 |
| 13 | Non-essential therapy ceased more than 24hrs prior to death? | 17 (15) | 11 (24) | 0.25 |
| 14 | Nursing care plan revised? | 83 (75) | 35 (80) | 0.69 |
| 15 | Opiate PRN order? | 82 (74) | 42 (91) | 0.02 |
| 16 | Benzodiazepine PRN order? | 48 (43) | 28 (61) | 0.05 |
| 17 | Antisialogogue PRN order? | 21 (19) | 12 (26) | 0.53 |
| 18 | Antiemetic PRN order? | 32 (29) | 20 (43) | 0.09 |
| 19 | LMO notified of death? | 71 (64) | 29 (63) | 1.00 |
| 20 | Bedcard Specialist notified of death? | 62 (56) | 33 (71) | 0.07 |
| 21 | Bereavement follow-up offered? | 30 (27) | 10 (22) | 0.55 |

We tested the DECoDE audit tool using retrospective chart review in two metropolitan university-affiliated hospitals, within the same health network, with a combined total of 2,000 ICU admissions and 200 ICU-related deaths per year. All records of adults (age ≥ 18-years) who died in ICU in the index hospital were eligible. Records were excluded if the death occurred after the patient was transferred from the ICU to another ward or hospital, since the primary aim was to measure care within the ICU. Decedents referred for organ donation after death and/or to the State Coroner were not excluded.

For the purpose of this report we also collected data on patient-specific factors present on admission to ICU, or during the last 24-hours of life, that may have influenced clinical decisions and end-of-life care. Admission data included age, sex, dignosis, and (Acute Physiology and Chronic Health Evaluation-IIIJ[20] [APACHE-3] model) severity of illness score and predicted risk of death (RoD) and chronic health burden.

Use of any life-support interventions in the last 24-hours were also recorded. These included mechanical ventilation (MV), renal replacement therapy (RRT), circulatory support with a vasopressor infusion (CVS), diagnostic imaging, and pathology testing. The time from redirection of care (withdrawal or withholding of curative therapy) to time of death, and the day of the week, were also recorded. Finally, the coding researcher made a judgement as to whether death was expected or not.

Patient records were randomly selected (by computer allocation) from those eligible (died while in ICU) during the 30-month period from January 2015 to June 2017. Study data were collected and stored using REDCap electronic data capture tools[21] hosted at the study hospitals. StataMP™ V15.1 (2017, College Station, TX) statistical software was used to analyse de-identified data.

Grouped data are reported as mean (and 95% confidence interval) or median (interquartile range) and results for the two sites were compared using Fisher’s exact test and Wilcoxon rank sum for categorical and continuous data, respectively. A random selection of 15 records were audited by another research member to assess interrater agreement (kappa concordance) of data extraction. Mixed effects regression was used to identify factors associated with DECoDe score. A P-value<0.05 was taken as significant. This research was approved by Eastern Health Research and Ethics Committee as a quality audit (QA97-2017) and the need for (third party) consent waived due to the observational nature of the audit.

## Results

Over the 30-month period there were 267,475 hospital and 5,194 (1.9%) ICU admissions. During the same time period there were 2,016 (0.8%) in-hospital deaths of which 358 (17.7%) occurred in ICU and 181 (9.0%) that occurred after transfer from ICU to another ward in the same hospital.

Compared to survivors, ICU decedents were significantly (P<0.001) older (74 years [95%CI = 64-81] vs 64 years [95%CI = 48-77]), had greater chronic health burden (2.2±4.6 vs 1.0±3.2) and higher APACHE-3 score (98 [95%CI = 74-124] vs 53 [95%CI = 38-70]) and predicted risk of death (0.60 [955CI = 0.30-0.81] vs 0.08 [95%CI = 0.3-0.20]).

Medical records of 157 (43.9%) randomly selected adults who died in ICU over a 30-month period (1 Jan 2015 – 30 June 2017) were reviewed by ICU junior medical staff who were independent of the treating team caring for the patient during the admission and at the time of death. The study population had higher severity of illness (APACHE score and RoD[20]) compared to the eligible population from which they were selected (P=0.04). There were no significant demographic differences between the cohorts from the two hospitals (Table 2).

**Table 2.** Demographics of study population. Data presented as median (interquartile range) unless otherwise specified. APACHE-3 = Acute Physiology and Chronic Health Evaluation version 3; RoD = risk of death; LOS = length of stay.

| Parameter | Site 1 | Site 2 | P-value |
| --- | --- | --- | --- |
| ICU admission, n | 2,889 | 2,305 | <0.001 |
| In-hospital ICU death, n (%) | 352 (12.2) | 187 (8.1) | <0.001 |
| Death in ICU, n (%) | 243 (8.4) | 115 (5.0) | <0.001 |
| Study population, n (%) | 111 (45.7) | 46 (40.0) | 0.36 |
| Age, years | 72 (65-80) | 73 (58-82) | 0.66 |
| Male, n (%) | 67 (60.4) | 24 (52.2) | 0.38 |
| APACHE-3 score | 110 (89-131) | 108 (85-139) | 0.75 |
| APACHE-3 RoD | 0.71 (0.49-0.86) | 0.68 (0.40-0.88) | 0.76 |
| LOS, days | 3 (1-7) | 3 (1-8) | 0.74 |
| DECoDE score | 11 (9-12) | 12 (9-14) | 0.06 |

A high proportion (n=129; 82.2%) of the study population were receiving major therapeutic interventions in the last 24-hours before death - 98(62.4%) were on vaso-pressors, 82 (52.2%) ventilated, and 23 (14.7%) receiving renal replacement (Table 3). There were 39 (25%) deaths referred to the State Coroner due to statutory requirements (without any adverse findings) and 11 (7%) were referred for organ donation. One hundred and thirty deaths (83%) were ‘expected’ and occurred after a median of 1.5 (1-4) days after family (or significant decision maker) assent to withdraw or withhold curative treatment and redirect care. These observations highlight some of the unique elements caring for the dying in an ICU.

**Table 3.** Study populations characteristics and patient management within the last 24-hours. Data presented as frequency (percentage) unless otherwise indicated.

| Patient Management | Site 1 | Site 2 | Combined | P-value |
| --- | --- | --- | --- | --- |
| Active Rx in last 24hrs | 94 (84.7) | 35 (76.1) | 129 (82.2) | 0.25 |
| Mechanical ventilation | 65 (58.6) | 17 (37.0) | 82 (52.2) | 0.02 |
| Vasopressors | 74 (66.7) | 24 (52.1) | 98 (62.4) | 0.10 |
| Antimicrobials | 56 (50.5) | 27 (58.7) | 83 (52.9) | 0.38 |
| Renal replacement | 20 (18.0) | 3 (6.5) | 23 (14.7) | 0.08 |
| Pathology testing | 86 (77.5) | 35 (76.1) | 121 (77.1) | 0.84 |
| Diagnostic Imaging | 65 (58.6) | 24 (52.2) | 89 (56.7) | 0.48 |
| Organ Donor | 7 (6) | 4 (9) | 11 (7) | 0.73 |
| Coronial referral | 29 (26) | 10 (22) | 39 (25) | 0.69 |
| Death 'unexpected' | 18 (16) | 6 (13) | 24 (15) | 0.81 |
| Withdrawal of care | 90 (81) | 40 (87) | 130 (83) | 0.49 |
| Time to Death, days (IQR) | 1 (1-4) days | 2 (0-4) | 1.5 (1-4) | 0.94 |

The median DECODE score was 11(IQR = 9-13) out of maximum possible score of 21. Kappa concordance between coders was 0.84. Factors associated with a higher DECoDE score included longer LOS, an ‘expected’ death, and a non-oncologic diagnosis (not shown). Patient factors such as age, severity of illness, or organ donation did not increase DECoDE score. There was no significant difference between DECoDE scores for each site.

## Discussion

We reviewed over 150 ICU deaths to test a simple tool - DECoDE - to screen the quality of documentation and care of dying patients in ICU. We found the tool simple to use, with high degree of concordance between staff, producing several important insights into patient care and future research.

While the DECoDE tool does not address all aspects of care it does incorporate major elements found in clinical guidelines promoted for the care of the dying patient.[12–15] The survey was designed as a ‘screening’ tool to direct further investigation rather than as ‘diagnostic’ test of the quality of care.

The DECoDE score has arbitrary units so the absolute score is difficult to interpret without repeated measures and comparison with an historical local benchmark. There is no ‘pass’ or ‘fail’ score. Comparison of scores between hospitals may be feasible but requires further validation and, possibly, adjustment for casemix.

A high DECoDE score does not necessarily equate with high quality documentation and care but a low score may identify gap(s) that warrant further investigation. For example, Question #7 (*Were the patient’s wishes documented?*) highlighted a potential gap in end-of-life care at both sites. Only a prospective investigation can, however, elucidate whether this was due to: (a) knowledge failure; (b) failure to ask; (c) failure to answer; or (d) failure to document.

Death in hospital is an uncommon event (0.8%) but 25% occur in ICU patients. We identified 2 to 5 cases per week in the study ICUs. Most of these deaths (90%) were ‘expected’. Not surprisingly we found that an ‘expected’ death, a longer LOS, and an oncologic diagnosis, were associated with a higher DECoDE score. Age, year and site of admission, day of week, and severity of illness scores were not significant.

Death in ICU occurred within a median of 72-hours of admission and 36-hours of re-direction of care. It is reassuring that these patients were given a trial of curative therapy and that their dying or suffering was not prolonged.[3] At the same time they raise concerns regarding the lack of time for families to prepare and staff to provide patient-centred high-quality end-of-life care.[4,5] Even though most of these patients were elderly with substantial comorbid disease it is possible that patient wishes and ‘goals of care’ were not discussed or known prior to admission.[9]

Future directions for research include repeat analysis following education and implementation of new guidelines or clinical pathways[12]. DECoDE may be applicable to other acute clinical services. Comparison of inter-and intra-hospital scores mandates adjustment for casemix and patient selection. Linkage with audit tools for the bereaved[23] are likely to add further insight and value to clinical audit.

As alluded to, there are several important limitations to the DECoDE tool. It requires manual chart review, does not identify cause, and comparison between hospitals and services is not recommended without further validation, and raw scores require clinical interpretation. Not all patients or services require a high score for high quality care, but a low score may highlight gaps in quality of care that warrant further review. In conclusion the DECoDE audit tool is simple to apply and permits screening and review of documentation and quality of care provided to dying patients. It may be useful as a baseline for assessment of future interventions to improve care and develop better audit tools.

## Data Availability

All data produced in the present study are available upon reasonable request to the corresponding author.

## Declarations

### Ethics approval and consent to participate

The Eastern Health Human Research Ethics Committee (QA97-2017) approved this study and patient consent was waived.

### Consent for publication

The Eastern Health Human Research Ethics Committee (QA97-2017) approved publication of this study. All authors approved this manuscript for publication.

### Author contributions

SD: methodology, data acquisition, data analysis, manuscript writing.

GD: concept, methodology, data acquisition, data analysis, manuscript writing.

JB: concept, methodology, manuscript writing.

SHu: data acquisition, data analysis, manuscript writing.

SHa: data acquisition, data analysis, manuscript writing.

DL: data acquisition, data analysis, manuscript writing.

NJ: data acquisition, data analysis, manuscript writing.

JS: data acquisition, data analysis, manuscript writing.

AT: data acquisition, data analysis, manuscript writing.

PZ: data acquisition, data analysis, manuscript writing.

## Acknowledgment

We thank the nursing staff who cared for the patients and families included in this study.

